# Performance-based assessments of cognition are less susceptible to demographic effects than traditional memory tests: Evidence from MindCrowd

**DOI:** 10.64898/2026.02.26.26347220

**Authors:** Alexandra M. Reed, Matthew J. Huentelman, Andrew Hooyman, Lee Ryan, Megan Johnson, Matt D. De Both, Saurabh Sharma, Darian Chambers, Matthew Calamia, Sydney Y. Schaefer

## Abstract

**Objective:** Demographic corrections (e.g., sex, education, race, ethnicity) are often applied when assessing cognition in adults; however, these corrections have significant limitations (e.g., using years of education does not capture the quality of, or access to, education). It is therefore critical to develop novel assessment options that are less susceptible to demographic factors. This study compared demographic effects on a verbal memory test and a performance-based test of cognition and daily functioning in older adults. Based on prior work, we hypothesized the performance-based tests would be less susceptible to demographic factors than paired associates learning.

**Method:** Data from 1326 participants (mean±SD age=61.9±10.9 yrs; Female = 1066, 80%) were collected through the MindCrowd electronic cohort, with 79 (6%) non-White, 109 (8.2%) identifying as Hispanic/Latino ethnicity, and 327 (25%) reporting education as less than a college degree. Paired associates learning is a well-established measure of medial temporal lobe-dependent learning and memory through recall of word-pairs, scored as the number of correct word pairs entered out of 36 possible. The performance-based test involved functional upper-extremity movement, specifically transporting beans to target cups in a repeating sequence (a task also shown to be dependent on the medial temporal lobe), scored as the intraindividual variability (standard deviation) in trial time across four consecutive trials.

**Results:** As hypothesized, linear regression analysis showed that PAL was significantly affected by sex, education, race (particularly Black/African American), and ethnicity, whereas the performance-based test was affected only by sex and with a much smaller effect size than that of PAL.

**Conclusions:** Performance-based assessments may be an equitable approach to evaluating cognition without requiring score corrections, particularly for diverse populations.

## INTRODUCTION

Numerous tests used for screening and evaluating cognition in older adults are influenced by demographic factors, such as education, race, and ethnicity. For example, higher education levels have been associated with better performance on the Mini-Mental State Examination (MMSE) (Crum & Folstein, 1993). Effects of ethnicity have also been shown for the Mini-Mental Status Exam (Bohnstedt et al., 1994), Montreal Cognitive Assessment (Milani et al., 2018), Clock Drawing Test (Menon et al., 2012), the Repeatable Battery for the Neuropsychological Assessment (RBANS) (Barnes-Marrero et al., 2022), the Brief Visuospatial Memory Test-Revised (Díaz-Santos et al., 2021), and the NIH Toolbox Cognition Battery (Flores et al., 2017). These effects can impact estimations of cognitive impairment in large-scale screenings (Edmonds et al., 2016), rates of inclusion/exclusion from research studies (including clinical trials), medication referral if a patient is considered too progressed (Jette et al., 2023; Xiong et al., 2015), and the overall ability to assess those from outside of majority groups (Byrd & Rivera-Mindt, 2022). As such, demographic corrections are often applied when assessing cognition in older adults; however, these corrections have significant limitations (e.g., using years of education does not capture the quality of, or access to, education). Additionally, the inclusion of factors such as race, which serves as a proxy for a host of other variables more proximally related to test performance, can be associated with harmful misinterpretations of test results (Manly, 2005). It is therefore critical to develop novel assessment options that are less susceptible to demographic factors.

Prior work has developed a brief (∼5-minute) performance-based test of cognition and daily functioning that correlates with performance on verbal memory tests such as the Repeatable Battery for the Assessment of Neuropsychological Status (RBANS) List Learning and List Recall and the Hopkins Verbal Learning Test-Revised (HVLT-R) Total and Delayed Recall, as well as global measures like the Mini-Mental Status Exam (Malek-Ahmadi et al., 2023). This test is also associated with putative markers of Alzheimer’s disease and neurodegeneration (e.g., beta-amyloid, hippocampal atrophy) and symptoms (i.e., cognitive and functional impairment) (Schaefer, Duff, et al., 2022; Schaefer, Malek-Ahmadi, et al., 2022; Schaefer et al., 2025), and is prognostic of cognitive decline (Koppelmans et al., 2025). Briefly, this performance-based test involves functional upper extremity movement in which the individual moves small objects with a spoon in a sequence with their nondominant hand. Prior work has shown that this performance-based test showed no significant effects of sex or education in a clinical sample of cognitively unimpaired, amnestic Mild Cognitive Impairment, and mild Alzheimer’s disease participants, whereas verbal memory tests did (as did other memory tests, such as the Brief Visuospatial Memory Test-Revised) (Reed et al., 2025).

However, this prior study was relatively small (<200 participants) and did not have sufficient variation across participant race and ethnicity to statistically evaluate these demographic effects. By leveraging a larger and more heterogeneous cohort, this study aimed to compare demographic effects on both a verbal memory test (paired associates learning) and the performance-based test in older adults. Based on prior work (Reed et al., 2025), we hypothesized the performance-based test would be less susceptible to demographic factors (sex, education, race, and ethnicity) than paired associates learning.

## METHODS

### Participants

This study utilized the MindCrowd electronic cohort (Huentelman, Talboom, Lewis, Chen, & Barnes, 2020). MindCrowd is an internet-based platform focused on neuroaging research that collects participant data via online surveys and assessments (Talboom et al., 2019). All participants in this study had no self-reported history of mental illness, Alzheimer’s disease, Parkinsons’s disease or stroke, and had no missing demographic data (age, sex, education, race, and ethnicity). All participants reported normal visual acuity and absence of any peripheral sensory or motor loss/pathology. Although MindCrowd itself has users worldwide, all participants in this study resided in the United States.

As of September 2025, a total of 1,326 participants with a reported age of 40 years or older (mean±SD age = 61.9±10.9 years) had completed the performance-based test (referred to as the ‘bean game’ within the MindCrowd study) in addition to the verbal memory test (described below), with no missing demographic data. Demographic data are summarized in Table 1. The WCG Institutional Review Board approved all portions of the study, with participants providing informed consent prior to enrolling.

**Table 1.** Summary of participants.

| **Total sample** | **N = 1,326** |
| --- | --- |
| Age | 61.9 (10.9) |
| Sex |  |
| Female | 1,066 (80%) |
| Male | 260 (20%) |
| Race |  |
| American Indian or Alaska Native | 3 (0.2%) |
| Asian | 22 (1.7%) |
| Black or African American | 18 (1.4%) |
| I prefer not to answer | 8 (0.6%) |
| More than one | 28 (2.1%) |
| White | 1,247 (94%) |
| Hispanic/Latino ethnicity (Yes) | 109 (8.2%) |
| Highest level of education |  |
| High school diploma (Baccalaureate) | 82 (6.2%) |
| Some college | 245 (18%) |
| College degree (University graduate) | 504 (38%) |
| Post-graduate degree (Masters or Doctorate) | 495 (37%) |
| Self-reported memory problems<sup>a</sup> | 191 (14%) |
| Paired-Associates Learning (total pairs correct)<sup>b</sup> | 21 (8) |
| Performance-based test score<sup>c</sup> | 6.8 (4.5) |

Mean (SD) or n (%). <sup>a</sup> Answered “yes” to the survey question “Have you personally experienced or are you currently experiencing memory problems?”; <sup>b</sup> out of 36 possible correct pairs; <sup>c</sup> scored as the intra-individual standard deviation in trial time across four consecutive timed trials, where lower is considered better.

### Assessments

#### Performance-based test

A visual description of the performance-based test can be viewed on Open Science Framework (https://osf.io/phs57/wiki/Functional_reaching_task/). To summarize, participants use a plastic spoon to acquire two raw kidney beans at a time from a central cup to one of three distal cups arranged at a radius of 16 cm at -40°, 0°, and 40° relative to the central cup. All cups are the same size (9.5cm diameter and 5.8cm deep). For the at-home version, individual kits were mailed to participants that had the four cups, 30 beans, the spoon, written instructions, and a paper ‘gameboard’ that provided a visual of where the home and target cups should be placed, along with 1” circular stickers to adhere the gameboard to a table in front of the participant when seated. More details about the at-home version of this test in MindCrowd have been published previously (Hooyman et al., 2021); the cost of materials and domestic shipping combined was <$10 per participant. Participants started acquiring two beans with the spoon and transporting them to the left target cup, then returned to the central cup to acquire two more beans to transport to the middle target cup, then the right target cup, and then repeated this 3-cup sequence five times for a total of 15 reaches. The trial ended once the last two beans were deposited into the last cup. Performance was measured as the amount of time it took to complete all 15 reaches, i.e., ‘trial time.’ Four trials were completed with the nondominant hand. Participants reported whether they timed themselves (70%) or were timed by a partner (30%). The test was scored for each participant based on the intraindividual variability (standard deviation) in trial time, where higher scores (i.e., more variability) are considered worse (Malek-Ahmadi et al., 2023; Malek-Ahmadi et al., 2025; Reed et al., 2024; Reed et al., 2025; Schaefer, Malek-Ahmadi, et al., 2022; Schaefer et al., 2025), consistent with interpretations of intraindividual variability in other neuropsychological assessments (Christ et al., 2018; Halliday et al., 2018; Holmqvist et al., 2022; Jutten et al., 2024; Kälin et al., 2014; Scott et al., 2023). This test and its scoring method have been shown previously to be significantly associated with the medial temporal lobe, both in terms of volume and thickness (Malek-Ahmadi et al., 2023; Malek-Ahmadi et al., 2025). As noted above, this test is referred to as the ‘bean game’ within MindCrowd.

#### Paired Associates Learning

Paired associates learning (PAL) is a classic memory paradigm from the Wechsler Memory Scale used in neuropsychology that has been shown to be medial temporal lobe-dependent (Eichenbaum & Bunsey, 1995; Pike et al., 2008; Rizzuto & Kahana, 2001), representing one of the most sensitive measures of memory impairment in both neurological and psychiatric populations (Han et al., 2017; Pike et al., 2013; Wilson et al., 1989). The PAL presents 12 word-pairs during the learning phase of a given trial, one word-pair at a time (2 seconds/word-pair) on the screen. During the recall phase, participants were presented with the first word of each pair and were asked to use their keyboard to type (i.e., recall) in the missing word. This learning-recall procedure was repeated for two additional trials, with each trial having 12 different word-pairs. Prior to beginning the task, each participant had one practice trial consisting of three word-pairs not contained in the 36 total pairs used during the test. Word-pairs were presented in different random orders during each learning and each recall phase. Participants were required to initiate a response within eight seconds, although there was no limit to response time as long as it was initiated within the time window. The dependent variable was the total number of correct word pairs entered across the three trials (i.e., 12 × 3 = 36, a perfect score).

### Statistical analyses

Two separate linear regression models were generated to examine the extent to which demographic factors (namely sex, education, race, and ethnicity) affected performance on the performance-based test (model 1) and paired associates learning (model 2). Independent variables for each model included age (in years), sex (male/female), highest level of education completed (High school diploma, Some college, College degree, Post-graduate degree), race (American Indian or Alaska Native, Asian, Black or African American, More than one race, White, or Prefer not to answer), and Hispanic/Latino origin (Yes/No). All analyses were performed in R version (4.0.3). Model assumptions of normality, heteroscedasticity, collinearity, and outliers were checked; scores on the performance-based test were log-transformed to meet the assumption of normality. Thus, for better interpretation, any significant beta coefficients in the regression model for the performance-based test (model 1) were back-transformed by taking their exponent and converted into the geometric, rather than arithmetic, mean. Estimated effect sizes were calculated for all significant categorical and continuous variables, with Cohen’s *d* values reported for categorical variables based on between group estimated marginal means, and partial R^2^ was reported for continuous variables.

## RESULTS

For the performance-based test (“bean game”), linear regression analysis showed that sex (β_exp(sexMale)_ = 1.09, 95% CI = [1.009; 1.19], *p* = .03, Cohen’s *d* = -.15) was the only significant effect on test scores (reported as the standard deviation in trial time). Specifically, males had 9% worse (higher) scores than females. As expected, test scores were dependent on participant age (β_exp(age)_ = 1.003, 95% CI = [1.0001, 1.006], *p* = .04, partial R^2^ = .003), such that every one-year increase in age was related to a 0.3% increase in test score. Thus, for interpretation the predicted bean game score for a 65-year-old male would be 5.81, whereas the predicted score for a 65-year-old female would be 5.29, approximately a 0.52 back-transformed difference in scores between males and females. The estimated effect size of the calculated sex differences was small (Cohen’s *d* = .15). Notably, bean game test scores were not significantly related to education level (F(3,1314) = .25, *p* = 0.86), race (F(5,1314) =0 .74, *p* = 0.59), nor ethnicity (F(1,1314) = 0.18, *p* = 0.67).

For paired associates learning, linear regression analysis showed that both sex (β_sexMale_ = −4.25, 95% CI = [−5.26, −3.24], *p* < .001, Cohen’s *d* = 0.58) and ethnicity (β_Hispanic/LatinoYes_ = −3.02, 95% CI = [−4.55, −1.49], *p* < .001, Cohen’s *d* = 0.41) significantly affected performance (scored as the number of correct pairs, out of 36). Specifically, males scored worse by 4.25 points than females, and participants who reported ethnicity as Hispanic/Latino origin scored worse by 3.02 points than those who reported being not Hispanic/Latino. The estimated effect size of the calculated sex differences was moderate (Cohen’s *d* = .58), approximately four times larger than that for the bean game. There was also a significant effect of education (F(3,1314) = 16.74, *p* < 0.001), as well as race (F(5,1314) = 2.50, *p* = .03). After correction for multiple comparisons (Tukey method), pairwise comparisons between race categories showed that participants who identified as white had scores 6.14 points better than those who identified as Black or African American (*adjusted p* = .007, Cohen’s *d* = -0.83). All other pairwise comparisons were not statistically significant. As expected, age was significantly negatively associated with PAL performance (β_age_ = −0.15, 95% CI = [−0.19, −0.11], *p* < .001, partial R^2^ = 0.05), indicating that older age was linked to poorer performance.

## DISCUSSION

This study used the MindCrowd cohort to compare demographic effects on a verbal memory test (paired associates learning) and a performance-based test of cognition and daily functioning in over 1,300 adults nationwide. Results showed only a small effect of sex on the performance-based test, with no other demographic effect (education, race, or ethnicity) being significant. In contrast, all effects of sex, education, race, and ethnicity were significant for paired associates learning, consistent with (Nuño et al., 2026; Ryan et al., 2025). By leveraging large samples, this study offers an alternative to traditional testing methods used within neuropsychology that require demographic corrections for interpretation, providing new approaches for screening and monitoring cognitive and functional status in a wider range of older adults.

While prior comparisons of demographic effects included in-person testing (Reed et al., 2025), this study utilized data collected remotely and unsupervised. Thus, this study provides further validation of the bean game in at-home settings, which may be a more appropriate environment for testing overall (Schlemmer & Desrichard, 2018), and the reliability of the remote, at-home version of the bean game has been established previously in a similar age range (Hooyman et al., 2021). As such, this study also suggests that the bean game could be used in teleneuropsychology (Bettcher et al., 2025; Butterbrod et al., 2024; Yildirim et al., 2025) to easily and affordably pre-screen for biomarker testing, predict cognitive decline (e.g., prognosis), and/or monitor treatment outcomes without needing to apply or consider demographic corrections. Remote testing can drastically reduce clinical trial costs (DiMasi et al., 2023; Fu et al., 2023) while also improving recruitment and adherence (Anderson et al., 2018; Rosbach & Andersen, 2017).

There were several limitations to this study. First, we acknowledge that the sample is not representative of the general population (see Table 1). It is plausible that in more diverse or representative samples, the performance-based test may show significant demographic effects; however, this study was sufficiently powered to detect demographic effects in paired associates learning, indicating the effect sizes of age, sex, education, race, and ethnicity are much larger than for the performance-based test. Future studies can consider cultures and populations outside of the United States to determine the extent to which the bean game and other assessments (traditional in-person testing or emerging digital biomarkers) (Polk et al., 2025) are affected by demographic variables. Second, we acknowledge that no extensive neuropsychological assessment was used to evaluate cognitive status in this convenience sample; thus, the extent to which participants were cognitively unimpaired or impaired remains unknown. Nevertheless, future studies in well-characterized clinical samples can better compare the extent to which cognitive status (e.g., Mild Cognitive Impairment vs. dementia) and demographic factors (e.g., race, ethnicity) interact when scoring neuropsychological assessments.

## Data Availability

All data produced in the present study are available upon reasonable request to the authors.

